# Rare Variants in Pharmacogenes Influence Clozapine Metabolism in Individuals with Schizophrenia

**DOI:** 10.1101/2023.03.13.23287157

**Authors:** Djenifer B. Kappel, Elliott Rees, Eilidh Fenner, Adrian King, John Jansen, Marinka Helthuis, Michael J. Owen, Michael C. O’Donovan, James T.R. Walters, Antonio F. Pardiñas

**Affiliations:** Centre for Neuropsychiatric Genetics and Genomics, Division of Psychological Medicine and Clinical Neurosciences, School of Medicine, Cardiff University, Cardiff, UK; Magna Laboratories Ltd., Ross-on-Wye, UK; Leyden Delta B.V., Nijmegen, the Netherlands

**Keywords:** schizophrenia, clozapine, metabolism, whole exome sequencing, pharmacogenomics

## Abstract

Clozapine is the only licensed medication for treatment-resistant schizophrenia (TRS). Few predictors for variation in response to clozapine have been identified, but clozapine metabolism is known to influence therapeutic response and the occurrence of adverse side effects. Here, we expand on genome-wide studies of clozapine metabolism, previously focused on common genetic variation, by analysing whole-exome sequencing data from 2062 individuals with schizophrenia taking clozapine in the UK. Our main aim is to investigate whether rare genomic variation in genes and gene sets involved in the clozapine metabolism pathway influences plasma concentrations of clozapine metabolites, assessed through the longitudinal analysis of 6585 pharmacokinetic assays. We observed a statistically significant association between the burden of rare damaging coding variants (MAF ≤1%) in gene sets broadly related to drug pharmacokinetics and lower clozapine (β= -0.054, SE= 0.019, P-value= 0.005) concentrations in plasma. We estimate that the effects in clozapine plasma concentrations of a single damaging allele in this gene set are akin to reducing the clozapine dose by about 35 mg/day. Gene-based analysis identified rare variants in *CYP1A2*, which encodes the enzyme responsible for converting clozapine to norclozapine, as having the strongest effects of any gene on clozapine metabolism (β= 0.324, SE= 0.124, P= 0.009). Our findings support the hypothesis that rare genetic variants in known drug-metabolising enzymes and transporters can markedly influence clozapine plasma concentrations. These results also converge with common variant evidence, particularly in relation to *CYP1A2*, suggesting the need for further evaluations of the pharmacogenomic makeup of this gene. Overall, our results suggest that pharmacogenomic efforts trying to predict clozapine metabolism and personalise drug therapy could benefit from the inclusion of rare damaging variants in pharmacogenes beyond those already identified and catalogued as PGx star alleles.

## Introduction

Schizophrenia is a chronic psychiatric disorder affecting 7 out of 1000 people worldwide ^1^. Between 20-30% of individuals with schizophrenia develop treatment-resistant schizophrenia (TRS), a particularly severe presentation of the disorder in which symptoms do not respond to treatment after at least two courses of standard antipsychotics^2, 3^. Clozapine is the first-line treatment for TRS, but its use is associated with adverse effects which require careful clinical management^4^. In particular, due to the risk of a haematological adverse drug reaction (ADR) – agranulocytosis – individuals treated with clozapine undergo regular mandatory haematological monitoring. This monitoring can also be used to ensure treatment adherence by measuring concentrations of the drug and its metabolites in plasma. Therapeutic Drug Monitoring (TDM) frameworks have also been proposed to ensure clozapine dosing leads to plasma concentrations within the standard “therapeutic range” (350-600 ng/mL) that aims to balance the likelihood of treatment response with the risk of dose-dependent ADRs^5, 6^. However, even in high-income countries with extensive public health systems, TDM is not always used by clinicians for several reasons including the low availability of testing, and a lack of familiarity with how to apply it in clinical decision-making^7^.

Drug pharmacokinetics can vary substantially between people, with both environmental and genetic factors partially explaining these differences. While 40% of the inter-individual variability in clozapine metabolism has been related to demographic, lifestyle and clinical predictors (sex, age, body weight and smoking)^8^, the few genetic studies performed to date have already found a small number of common genetic variants explaining up to an extra 3%-8%^9^. As the effects of these markers are additive to those of environmental predictors^10^, their discovery is a first step towards eventual pre-emptive pharmacogenomic (PGx) interventions to personalise and improve clozapine treatment protocols.

Most PGx studies of clozapine and other psychiatric medications have been based on the “candidate study” experimental design, investigating well-characterized common polymorphisms in genes encoding cytochrome P450 enzymes (CYPs) that in many cases have been mapped to haplotypes commonly known as “star alleles”^11^. These PGx markers are routinely assessed by genetic testing companies and, given their strong effects on enzyme activity^12^, they have been proposed as useful to guide drug selection and dosing. The hepatic first-pass metabolism of clozapine is driven by CYP1A2 (with a minor role played by CYP3A4 and CYP2D6), and therefore lifestyle factors and concomitant medications influencing the activity of this enzyme are commonly discussed in the literature^6^. However, a clear relationship between *CYP1A2* genetic variation and clozapine metabolism has not yet been established, and no genetic variants are considered relevant for clinical management on recent expert reviews^13, 14^ or in the PharmGKB database^15^. The star allele *CYP1A2*\*1F has been the most consistently studied polymorphism as it is indexed by a single common SNP (rs762551; global minor allele frequency [MAF] 38%^16^) and leads to ultra-rapid metabolism in conjunction with enzyme inducers^17^. Despite multiple candidate studies likely sparked by its proposed relevance for other metabolites and conditions^18^, there is little evidence for *CYP1A2*\*1F association with clozapine pharmacokinetics or response^19-21^. Genome-wide association studies (GWAS) of clozapine plasma concentrations have also not implicated variants within the *CYP1A2* gene, although a 13kb upstream variant potentially involved in its regulation, rs2472297, has a reported and replicated association^10, 22^. It is important to consider that both GWAS and candidate gene studies are best suited to identify the influences of relatively common alleles. Sequencing studies are required to detect rare genetic variation, which is responsible for most of the known variability of drug-related enzymes and transporters (“pharmacogenes”)^23, 24^. Rare variants with a functional effect on pharmacogenes can thus become consequential for a given carrier when a relevant drug is prescribed, and thereby contribute to individual-level variability in drug metabolism and ultimately become clinically actionable through downstream effects on medication response, tolerability, and other outcomes^17, 25^.

In this study, we sought to complement the growing literature on candidate studies and GWAS of clozapine metabolism by assessing whole exome sequencing (WES) data from over 2000 individuals taking this drug. Our aim was to investigate for the first time, whether rare variants (MAF ≤1%) in genes involved in the clozapine metabolism pathway affect clozapine pharmacokinetics.

## Subjects and Methods

### Participants

Participants were from the CLOZUK2 study^26^ which received UK National Research Ethics Service approval (ref. 10/WSE02/15) in accordance with the UK Human Tissue Act. As previously reported^22, 26^, CLOZUK2 includes clozapine monitoring records from the Zaponex® Treatment Access System (ZTAS) and genetic information from anonymised individuals with TRS taking clozapine in the United Kingdom. Pharmacokinetic data derived from the monitoring records consisted of clozapine and norclozapine plasma concentrations (also known as “levels”) retrieved during 2013-2015. Briefly, data curation was performed as described in another study^27^, ensuring that all pharmacokinetic assays included in our analyses followed the following criteria: individual at least 18 years old and taking clozapine doses ≤ 900 mg/day; blood was drawn at an interval of 6 to 24 hours after their last clozapine dose; clozapine levels not exceeding 2000 ng/mL. The final curated dataset contained 3578 individuals linked to 11407 pharmacokinetic assays.

### Whole Exome Sequencing

Whole exome sequencing data was retrieved for 2405 CLOZUK2 participants linked to curated pharmacokinetic information. Sequencing data were generated using the Nextera DNA Exome capture kit and the Illumina HiSeq 3000/4000 platform (Illumina Inc, California), as previously described^28^. Raw sequence data were processed following the Genome Analysis Toolkit (GATK) best practices pipeline ^29^, and aligned to the human reference genome (GRCh37) using the Burrows–Wheeler Aligner (bwa) v0.7.15^30^. Variants were jointly called using GATK haplotype caller (v3.4) and filtered using the GATK variant quality score recalibration tool, VQSR. Processed sequencing data were queried using Hail^31^ and standard quality control procedures were performed, excluding samples where one or more of the following conditions was true: a) less than 70% of the exome target achieved 10× coverage; b) mismatch between genetically inferred sex and recorded sex; c) presence of another sample with a second degree or closer genetic relationship; d) failed sequencing quality control hard filter. Further details about the sequencing procedure and quality control are presented in the **Supplementary Information**. After this process, 2062 high-quality exomes remained for further analyses.

### Variant and Gene Set selection

We focused on variants with a MAF ≤1% in both our sample and the gnomAD control cohorts^32^. From those, we selected three classes of functional variants: synonymous variants, protein truncating variants (PTVs – frameshift, splice, start-lost, stop-gain), and damaging missense variants with a CADD^33^ PHRED-score above 20. The latter is a threshold optimised for pharmacogenes^34^, and both PTVS and damaging missense classes enrich for variants with a large impact on protein function^23^.

Variants fulfilling these criteria were grouped into genes and subsequently into pathways or gene sets of interest. As only a relatively small number of genes have evidence of direct roles in clozapine metabolism^25^, we also assessed gene sets relevant to broadly defined biological processes (absorption, distribution, metabolism and excretion; ADME) related to drug pharmacokinetics^35^. Four gene sets were selected *a priori* for analysis: the PharmGKB clozapine pathway set, including genes with consensus evidence of involvement in clozapine pharmacokinetics (15 genes); the PharmaADME core set, including genes related to general drug metabolism (33 genes); the PharmaADME extended set, incorporating drug targets (295 genes); and the CPIC Very Important Pharmacogenes (VIP) set, a curated list of genes used to derive pharmacogenomic recommendations for many medications (57 genes). **Supplementary Table 1** details the genes included in each of the gene sets. Note that as genes in these sets are involved in similar drug metabolism routes, there is a large degree of overlap, and their union represents 321 unique genes (**Supplementary Figure 3**).

### Statistical Analysis

All statistical analyses were conducted in R (v4.2.2). Rare variants located in genes included in the sets listed above were counted and aggregated in a burden metric within each of the four gene sets. This strategy is considered effective for jointly testing multiple rare variants, improving statistical power for association discovery. Generalised linear mixed-effect model (GLMM) regressions were used to assess the relationships between rare set-based burdens and plasma concentrations of the active parent compound (clozapine) and the main metabolite (norclozapine). Models were fitted using the *glmmTMB* package^36^ in R, which allows for defining fixed-effect predictors or covariates for both the outcome mean and variance. Covariates for the mean included in the models were: clozapine dose, the time between the last clozapine dose and blood sample collection (TDS), sex, age, age^2^, 10 genomic principal components derived from the exome sequencing data, 4 index SNPs previously identified in a clozapine metabolite GWAS of overlapping CLOZUK2 samples (rs1126545_T, rs2472297_T, rs61750900_T, rs2011425_G)^22^, and the burden of synonymous variants across the exome at the same MAF threshold. Covariates for the variance included: clozapine dose, TDS, sex, age, and age^2^. One random effect predictor per study participant was included to account for the longitudinal nature of the data and prevent confounding due to non-independence of the plasma concentrations within individuals. Further details about the statistical modelling procedure are given in the **Supplementary Note**.

Given the high overlap in the genes included in the four gene sets tested, as well as in the variant categories (PTVs, missense or both - PTVs + missense), the false discovery rate (FDR) using the Benjamini-Hochberg method was used as a correction for multiple testing at a p_FDR_≤0.1 threshold instead of the overly conservative Bonferroni procedure^37^. In each regression model, we estimated the variance explained by the rare variant burden using the semi-partial R^2^ method implemented in the *partR2* R package^38^. As this method is not implemented for the *glmmTMB* models described above, standard GLMMs were fitted using the *lme4* framework with the same fixed- and random-effect predictors used for this procedure (**Supplementary Note**).

Since the set-based burden used in the analyses was calculated across several genes, GLMM regressions with the same method and covariates as the primary analyses were fitted in a gene-based approach in order to disentangle gene-specific effects from the overall set-based associations. In this step, individual genes carrying fewer than 5 damaging variants were not analysed due to lack of power and increased likelihood of false positives. Multiple testing was accounted for using a Bonferroni correction corresponding to the number of genes being evaluated (α = 0.05/313).

## Results

After phenotypic and genetic quality control, 2062 individuals linked to 6585 clozapine plasma concentration assays (1-42 observations per individual) were retained: 72.8% were male (1501 male; 561 female), and the mean age was 43.61 years (SD= 11.38) at the last available clozapine monitoring measure. Over 6000 rare damaging nonsynonymous alleles (MAF ≤ 1%) affecting 313 of 321 genes included in the four gene sets were identified (624 PTVs; 5435 missense SNVs).

### Gene set Association Analyses

We observed significant associations between the burden of rare damaging mutations in the PharmaADME core set and both clozapine and norclozapine plasma concentrations (**Figure 1**). The combined PTVs + missense variant set included 837 rare damaging alleles affecting 29 genes in the core PharmaADME set. Each rare allele counted in this gene set was associated with lower clozapine (β= -0.054, SE= 0.019, P-value= 0.005) and norclozapine levels (β= -0.043, SE= 0.018, P-value= 0.015) when compared to individuals carrying zero damaging alleles, though only the clozapine association remained significant after FDR correction. Through comparison with other predictors in the statistical model (**Table 2**), we estimate that the effects in clozapine plasma concentrations of a single damaging allele in this gene set are akin to reducing the clozapine dose by about 35 mg/day (**Figure 2**). **Supplementary Table 2** details the main results for all gene sets for both clozapine and norclozapine levels, including the percentage of pharmacokinetic variance explained by rare variants in these sets.

**Figure 1:**
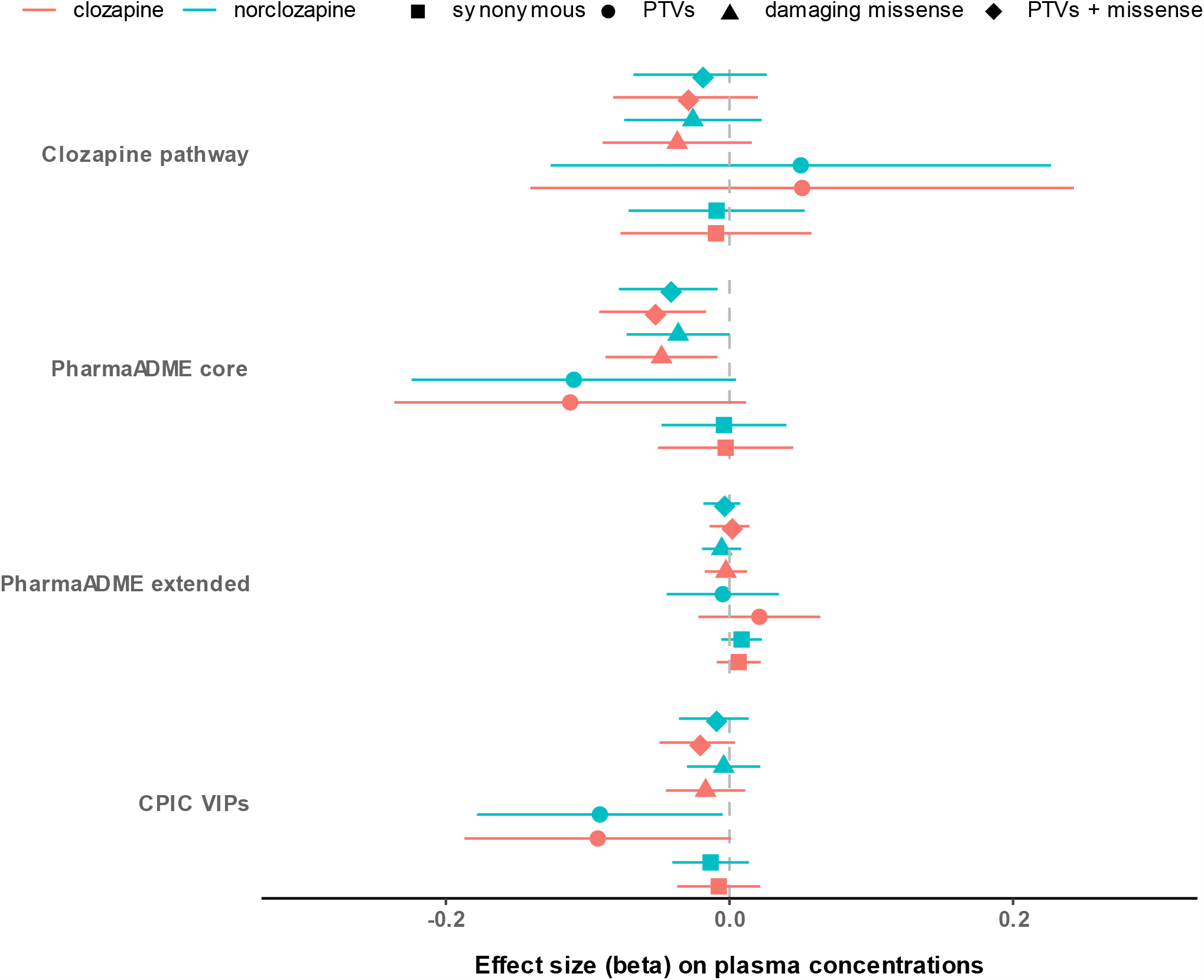
Estimated effect sizes and confidence intervals for the different variant classes in each of the four gene sets analysed. Supplementary Table 2 gives the full results for all gene sets for both clozapine and norclozapine levels, including the percentage of variability explained by rare variants in each of the sets.

**Figure 2:**
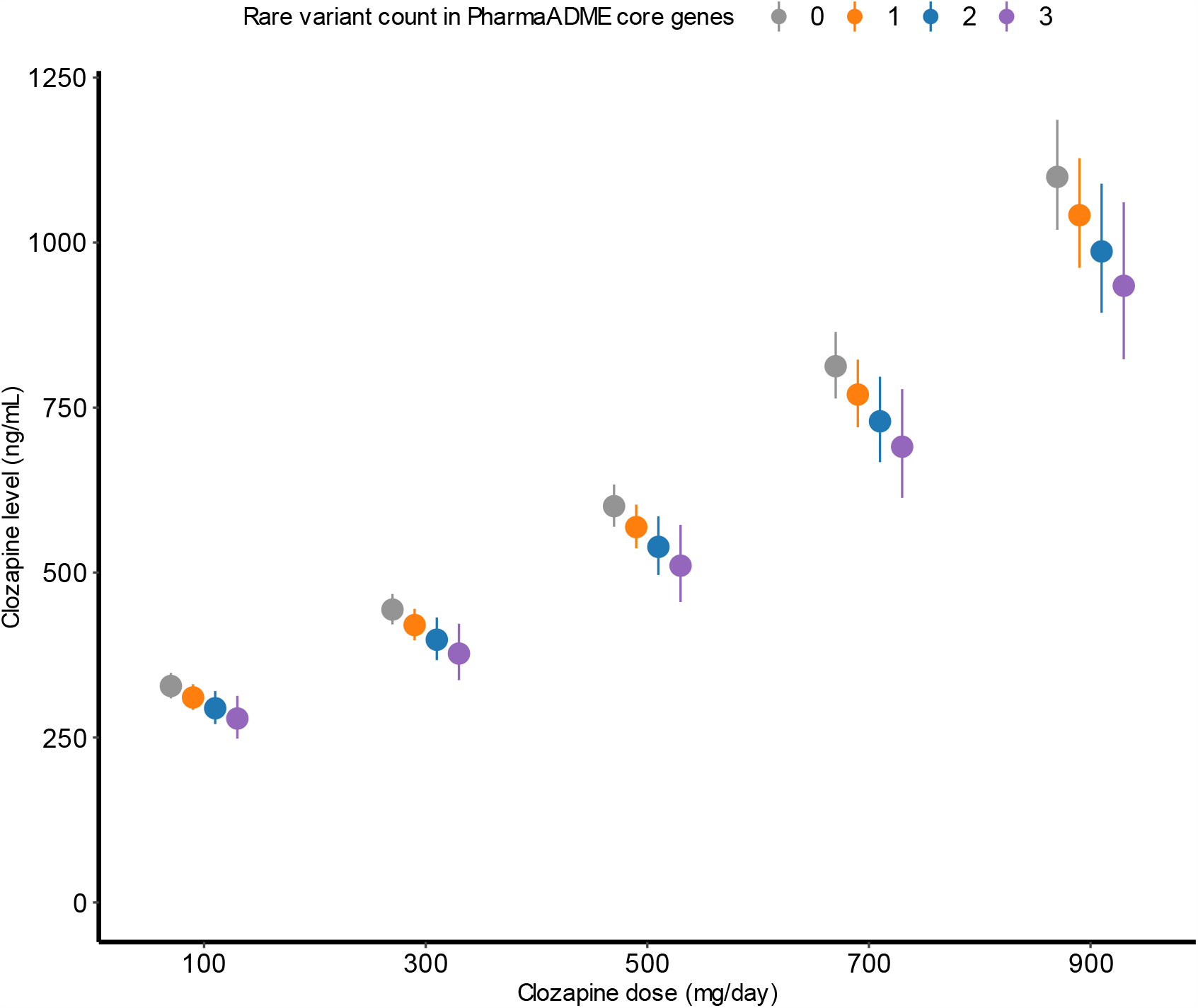
Marginal effects for individuals carrying different numbers of rare damaging variants in the PharmaADME core gene set in the relationship between clozapine doses and plasma concentrations. Dots and whiskers indicate the estimated value of the regression effect size and its corresponding 95% confidence interval at specific doses.

### Gene-Based Association Analyses

Next, we conducted a follow-up gene-based burden analysis focused on the genes within the PharmaADME core set (see **Table 1**). In total, 26 genes were included in this step, and 3 were excluded due to having fewer than 5 rare damaging nonsynonymous variants. No single gene was significantly associated with clozapine or norclozapine plasma concentrations after multiple testing corrections (**Supplementary Tables 3 and 4)**. We observed that rare damaging variants in *CYP1A2* displayed the strongest nominally significant associations with higher clozapine (β= 0.324, SE= 0.124, P= 0.009) and norclozapine (β= 0.320, SE= 0.115, P= 0.005) concentrations. Other nominal gene-level associations observed were *UGT1A1* with clozapine levels (β= -0.258, SE= 0.122, P= 0.034) and *SULT1A1* with norclozapine (β= 0.281, SE= 0.137, P= 0.041).

**Table 1:** Genes included in the PharmaADME core gene list

|  |  |
| --- | --- |
| PharmaADME<br>core | *ABCB1, ABCC2, ABCG2, CYP1A1, CYP1A2, CYP2A6, CYP2B6, CYP2C19, CYP2C8,*<br>*CYP2C9, CYP2D6, CYP2E1, CYP3A4, CYP3A5, DPYD, GSTM1, GSTP1, GSTT1, SLC6A2,*<br>*NAT2, SLC15A2, SLC22A1, SLC22A2, SLC22A6, SLCO1B1, SLCO1B3, SULT1A1,*<br>*TPMT, UGT1A1, UGT2B15, UGT2B17, UGT2B7* |

**Table 2:** Effect sizes for each of the genetic, demographic, and clinical covariates included in the GLMM in the PharmaADME core gene set association with clozapine plasma concentrations

| Predictor | $\beta$ | SE | P | Component |
| --- | --- | --- | --- | --- |
| PTVs+missense in PharmaADME core set | -0.0542 | 0.0193 | 4.88E-03 | mean |
| Daily dose (mg/day) | 0.0015 | 0.0001 | 3.12E-176 | mean |
| Time between dose and blood draw (hours) ‡ | -0.0131 | 0.0059 | 2.57E-02 | mean |
| Sex (male) | -0.1108 | 0.0265 | 2.92E-05 | mean |
| Age (years) ‡ | 0.0416 | 0.0126 | 1.01E-03 | mean |
| Age<sup>2</sup> (years<sup>2</sup>) ‡ | 0.0098 | 0.0112 | 3.80E-01 | mean |
| rs1126545_T | 0.0233 | 0.0238 | 3.26E-01 | mean |
| rs2472297_T | -0.0984 | 0.0186 | 1.27E-07 | mean |
| rs61750900_T | 0.0311 | 0.0278 | 2.64E-01 | mean |
| rs2011425_G | -0.1149 | 0.0300 | 1.31E-04 | mean |
| PC1 ‡ | 0.0292 | 0.0136 | 3.18E-02 | mean |
| PC2 ‡ | 0.0131 | 0.0115 | 2.57E-01 | mean |
| PC3 ‡ | -0.0015 | 0.0116 | 8.95E-01 | mean |
| PC4 ‡ | -0.0051 | 0.0115 | 6.56E-01 | mean |
| PC5 ‡ | -0.0012 | 0.0117 | 9.19E-01 | mean |
| PC6 ‡ | 0.0032 | 0.0114 | 7.81E-01 | mean |
| PC7 ‡ | -0.0082 | 0.0115 | 4.74E-01 | mean |
| PC8 ‡ | -0.0052 | 0.0117 | 6.55E-01 | mean |
| PC9 ‡ | -0.0065 | 0.0114 | 5.71E-01 | mean |
| PC10 ‡ | -0.0022 | 0.0117 | 8.52E-01 | mean |
| Synonymous variant count | -0.0001 | 0.0007 | 8.83E-01 | mean |
| Daily dose (mg/day) | 1.9444 | 0.0632 | 4.89E-208 | variance |
| Time between dose and blood draw (hours) ‡ | 0.0008 | 0.0001 | 1.54E-09 | variance |
| Sex (male) | -0.0741 | 0.0218 | 6.71E-04 | variance |
| Age (years) ‡ | -0.1213 | 0.0489 | 1.31E-02 | variance |
| Age<sup>2</sup> (years<sup>2</sup>) ‡ | 0.0447 | 0.0222 | 4.42E-02 | variance |

Effect size coefficients ( $\beta$ ) indicate the positive or negative impact of a one-unit increase of the predictor in the average of clozapine concentrations in ng/mL. Predictors marked with ‡ were standardised to improve model fit and their effect sizes refer to a one standard deviation increase<sup>54</sup>.

**Table 3:** Estimated variance of clozapine plasma concentration explained by different sources of rare variants.

| Predictor | R<sup>2</sup> |
| --- | --- |
| Full model (see Table 2) | 0.18100 |
| PharmaADME core gene set | 0.00407 |
| *CYP1A2* | 0.00233 |
| PharmaADME core gene set (-*CYP1A2*) | 0.00503 |
| *CYP1A2* + PharmaADME core gene set (-*CYP1A2*) | 0.00736 |

### Leave-one-gene-out set association analyses

Rare alleles in *CYP1A2* variants had an opposite direction of effect to rare alleles in the full PharmaADME core set (**Figure 3 – Supplementary Table 5**), that is, they were associated with higher clozapine and norclozapine levels. Thus, we performed a sensitivity analysis by excluding *CYP1A2* from the PharmaADME set, recalculating the burden scores, and conducting the GLMM regression again. This yielded the same result of rare variant burden in the PharmaADME core set being associated with clozapine and norclozapine metabolism, and as expected the effect sizes were slightly stronger after the removal of *CYP1A2* (**Supplementary Figure 4**). We also tested for additive effects of separately considering rare damaging variants in *CYP1A2* and the remainder of the PharmaADME gene set, showing that the addition of both burdens as individual predictors resulted in an 80.83% increase in the variance of clozapine levels explained by rare genetic variation (**Table 3**). We did not find any evidence of multiplicative (i.e., non-linear) relationships between these two predictors in our dataset (interaction test P= 0.470).

**Figure 3:**
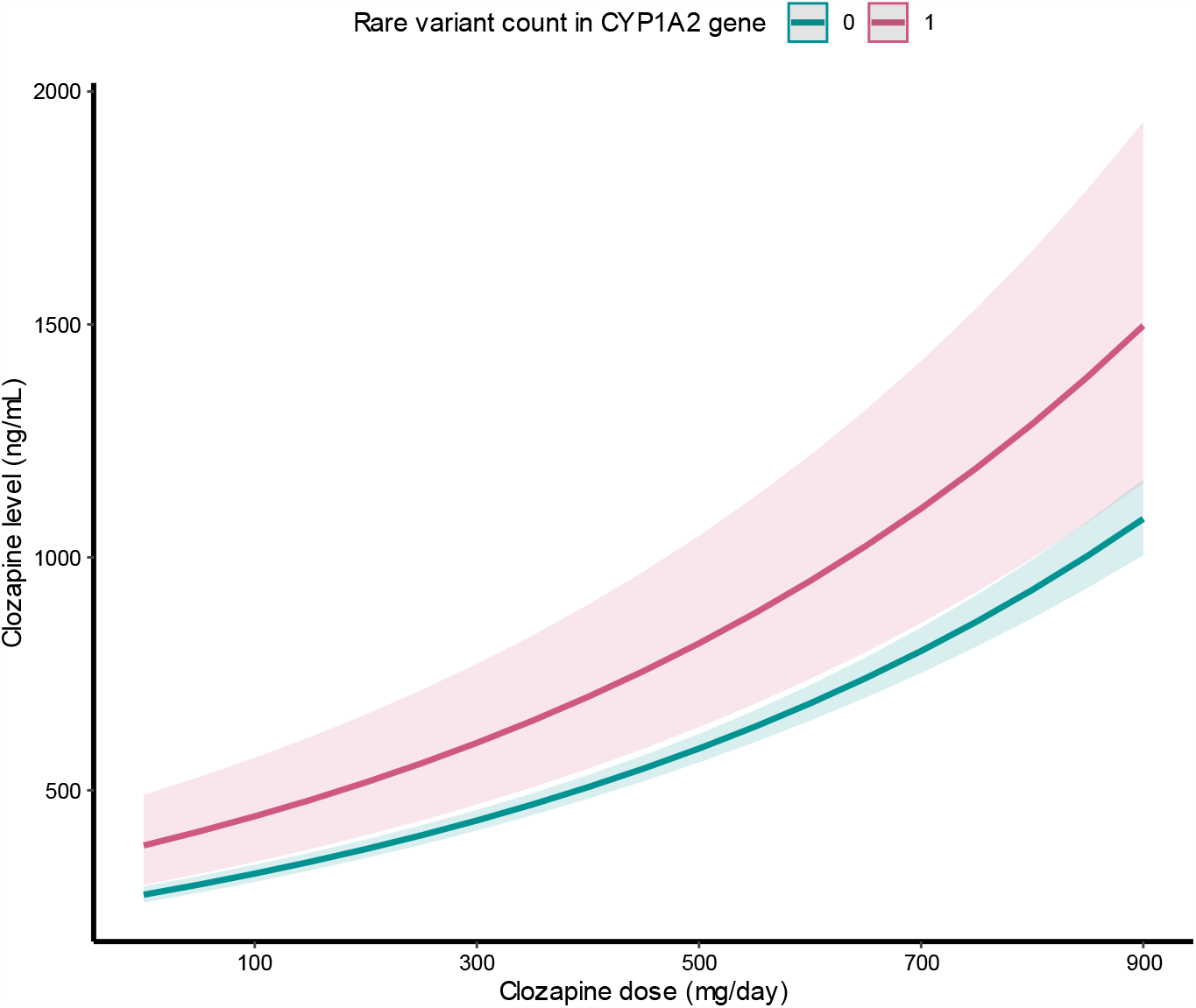
Marginal effects representing the relationship between clozapine doses and plasma concentrations in individuals carrying or not rare damaging variants in the *CYP1A2* gene. Shaded areas highlight a 95% confidence interval for the estimates.

## Discussion

To our knowledge, this is the largest sequencing study seeking to evaluate the effects of rare pharmacogenomic variants in clozapine metabolism. Using longitudinal clozapine monitoring data, we evaluated exome data obtained by WES to discover genes and gene sets harbouring rare variants associated pharmacokinetic variation. We provide evidence that deleterious variants in pharmacogenes can significantly affect clozapine metabolism, leading to different metabolite plasma concentrations for individuals prescribed the same drug dose.

Our primary analysis show that a greater burden of rare damaging variants in the PharmaADME core gene set, which contained 30 genes associated with general drug metabolism (**Table 1**), was associated with significantly lower clozapine and norclozapine plasma concentrations (**Figure 1**). Furthermore, the burden of rare variants in this set explains 0.40% of the variability of this phenotype (**Table 3**), a similar figure to the 0.61% variance explained by a PRS generated from genome-wide significant loci in a recent clozapine metabolism GWAS^9^. In the CLOZUK2 sample, the effects of each rare damaging allele in this set were approximately equivalent to a 35 mg reduction in clozapine daily dose. As our models assume a linear additive effect for the mutation burden, carriers of multiple rare alleles (the maximum seen in our study was 3) would require further dose adjustments to reach the expected levels of the wider population of non-carriers (**Figure 2**). Furthermore, through ordinal mixed-model regression analysis (**Supplementary Results**), we also observed that carriers of at least one damaging allele in the PharmaADME core set were 30% less likely than non-carriers in CLOZUK2 to reach the therapeutic range (350–600 ng/mL) of plasma concentrations of clozapine (OR= 0.694; SE= 0.145; P= 0.012; **Supplementary Figure 5**). Metabolic networks have properties that make them resistant to deactivation by deleterious genetic variation^39^, but this also makes them more likely to tolerate protein-damaging variants. Our findings suggest that individuals accumulating several rare damaging variants (as assessed in the gene set burden scores) might then have impaired biochemical functions relevant to drug response or even downstream disease risk^40^. However, we note this study is not powered to assess the strength of specific rare variants, which could also have different directions of effect for a given phenotype depending on their protein consequence.

In further analyses, we analysed the burden of rare variants within specific genes included in the PharmaADME core gene set. Although no single gene was significantly associated with clozapine or norclozapine plasma levels after Bonferroni correction, the most significant gene in this analysis was *CYP1A2* (P= 0.009). However, surprisingly, individuals carrying rare variants in *CYP1A2* showed higher plasma clozapine levels when taking the same clozapine dose as non-carriers (**Figure 3**). This was contrary to the PharmaADME core gene set results and prompted us to test for individual contributions to clozapine metabolism of rare variants within and outside *CYP1A2* (**Table 3**). *CYP1A2* encodes the main catalyst enzyme of the clozapine to norclozapine biotransformation, and while common variants in its proximity have been previously associated with clozapine metabolite levels^27, 41^, no such association yet exists for coding SNPs. Our results suggest that *CYP1A2* rare coding variants, not included in the common haplotypes and star alleles frequently assessed by candidate studies, might also influence clozapine metabolism. Further investigation will be needed to identify specific variants and map them to new PGx star alleles, if possible, as well as to understand if these results could potentially support initiatives for pharmacogenetic testing as a decision-support tool for clozapine clinics.

Although genetic factors that account for a significant proportion of the interindividual variability in antipsychotic drug metabolism have been suggested as useful to improve therapeutic response and avoid ADRs ^42^, an effective model for the clinical implementation of this information in guiding treatment choices is not yet available. This study supports the view that such a precision psychiatry framework might benefit from taking account of genetic variation beyond the traditional PGx star allele definitions assessed by commercial genetic testing companies^23, 27^. Our analyses included rare damaging variants affecting pharmacogenes based on in-silico functional approaches, and thus most of them have not yet been functionally assessed (**Supplementary Table 6**). Nevertheless, and while specific variants should be externally validated before being evaluated further, the effects we detect for the mutational burdens are of a sufficient magnitude to be relevant for particularly vulnerable subsets of the population of those on clozapine, such as people initiating treatment (when ADRs are most common^43^) or who only tolerate low drug doses. This knowledge, added to that coming from common variant analyses, might one day be incorporated into clozapine prescribing guidelines that at present rely on non-genetic characteristics to personalise dosing^14^. In the meantime, proactive therapeutic drug monitoring strategies remain a tried-and-tested approach for navigating the large inter-individual variability of clozapine metabolism and achieving beneficial clinical outcomes^6, 44^.

One limitation of this study is that rare variant analyses require a large sample size to allow the identification of single rare variants that are associated with complex traits. Although we had access to WES data, we chose to focus our efforts on a relatively small number of genes with robust *a priori* evidence of their involvement in drug metabolism, attempting to avoid limiting the power of our statistical tests through multiple testing. This strategy followed the rationale and results from other genomic studies of drug metabolism, showing that these traits are likely influenced by a small number of variants in relevant genes, each encompassing a relatively large effect^27, 45^. However, unlike candidate studies, which often have included variants with experimentally derived functional activity information, we selected variants based on existing protein impact prediction algorithms. This is a PGx-informed approach to variant prioritisation, selecting rare variants putatively affecting enzyme and transporter function^34, 46^, but can still include markers with little or no impact on gene product while omitting other potentially important variants. Thus, assessing and collecting more information on the functional genetic makeup of pharmacogenes, through either bioinformatics or direct experimental approaches^47, 48^, will be valuable to update these analyses and for further research into developing PGx biomarkers for metabolite pharmacokinetics and drug response.

Another limitation is that, as with most statistical testing of burden scores, the regression models fitted for gene- and set-based analyses in the GLMM framework assume that all rare variants under testing follow a similar direction of effects^49^. This is known not to be the case in disease studies (i.e., both risk-increasing and protective variants can exist) and is likely a simplification for metabolism as well. While, again, our sample size limits the testing we can reliably perform for individual genes, results from gene-based analyses, particularly for *CYP1A2*, suggest that future studies might benefit from statistical methods allowing for testing variants with multiple directions of effect.

Finally, due to the limited data collection performed for CLOZUK2, important environmental factors, such as cigarette smoking or the use of other medications, were not available to be included in our analyses. These are known to be responsible for a fraction of the inter-individual differences in clozapine metabolism and thus might mediate some of our results. Nevertheless, some of the covariates already included in our regression models might capture some of the variance due to these exposures. For example, previous studies have found that sex can be a partial marker of oral contraceptive use and smoking, both interactors of CYP1A2, capturing some of their effects in drug metabolism^50, 51^. A related issue is that while the CLOZUK2 cohort is derived from the diverse current population of the UK, the majority of individuals for which both WES and clozapine levels monitoring data were available were of European ancestry. Given that individuals of other biogeographical ancestries are known to have different clozapine pharmacokinetic profiles^9^, our study cannot assess a potential rare variant contribution to those known ancestral differences, and its results might not be straightforwardly transferrable to non-European populations. We note, however, that consistency of variant effect sizes across populations is a reasonable assumption for pharmacogenes^52^ and that the occurrence of deleterious variation in these gene sets is frequent (though individually rare) across continental ancestries^53^. Thus, the association between rare damaging burden and clozapine metabolism identified here could be of relevance beyond the European sample in which it was discovered.

In conclusion, we found that the burden of rare damaging variants within genes in drug metabolism pathways was associated with differences in clozapine metabolite plasma concentrations, explaining part of the genetic variability in clozapine metabolism phenotypes. Our results suggest that pharmacogenomic efforts trying to predict clozapine metabolism and personalise drug therapy could benefit from the inclusion of rare damaging variants in pharmacogenes beyond those already identified and catalogued as PGx star alleles. Further investigation is warranted to confirm and replicate these findings before they can be evaluated in prospective trials or considered in clinical settings.

## Data Availability

To comply with the ethical and regulatory framework of the CLOZUK project, access to individual-level data requires a collaboration agreement with Cardiff University. Requests to access deidentified datasets, data dictionaries, and other summaries from the CLOZUK project should be directed to James T R Walters.

## Acknowledgements

DBK and AFP were supported by the Academy of Medical Sciences “Springboard” award (SBF005\1083). The CLOZUK study was supported by grants from the Medical Research Council to Cardiff University: Centre (MR/L010305/1), Program (MR/P005748/1), and Project (MR/L011794/1, MC_PC_17212); as well as the European Union’s Seventh Framework program (279227, “CRESTAR”). ER is supported by a UKRI Future Leaders Fellowship Grant (MR/T018712/1). EF is funded by a Wellcome Trust Integrative Neuroscience PhD Studentship. We also acknowledge the support of the Supercomputing Wales project, partly funded by the European Regional Development Fund (ERDF) via the Welsh Government. We thank Joanne Morgan, Lesley Bates, Catherine Bresner and Lucinda Hopkins (Cardiff University) for laboratory sample management, Andy Walker (Magna Laboratories), Anoushka Colson (Leyden Delta) and Hreinn Stefansson (deCODE Genetics) for contributing to the sample collection, anonymisation, data preparation and genotyping efforts of the CLOZUK2 sample.

