## Supplementary Note for "Rare Variants in Pharmacogenes Influence Clozapine Metabolism in Individuals with Schizophrenia"

### Supplementary Methods

#### Exome capture

Samples were prepared for whole exome sequencing (WES) using the Illumina HiSeq 3000/4000 capture kits according to the manufacturer's protocol. Once prepared, the exome-captured library was then sequenced in the Illumina HiSeq platform using the paired-end method. Exome sequences had a median of 83% of all targeted bases covered at ≥10×, and samples were excluded if less than 70% of the exome target achieved 10× coverage. Raw data was processed to remove adaptors and low-quality reads, then aligned to the GRCh37 human reference genome with Burrows–Wheeler Aligner (bwa) v0.7.15[1]. Genome Analysis Toolkit (GATK) v3.4[2] was then used for recalibration of base quality scores, realignment around indels and variant calling (HaplotypeCaller).

#### Variant quality control

We then proceeded with the variant and genotype quality control procedures in Hail[3]. Initial processing removed variants failing the GATK[2] Variant Quality Score Recalibration (VQSR). The genotypes in the remaining variants were then filtered for depth (DP) ≥ 10, genotype quality score (GQ) ≥ 30, allelic balance (AB) < 0.1 in homozygous calls for the reference allele, AB ≥ 0.25 and ≤ 0.75 for heterozygous calls and AB ≥ 0.9 for homozygous calls for the alternative allele. Variants were also excluded if their call rate was < 0.97 or had a Hardy–Weinberg Equilibrium exact test P value of $< 1 \times 10^{-6}$.

#### Sex check

Genetic sex was inferred using a set of high-quality common variants on the X and Y chromosomes and from the rates of heterozygous and homozygous calls on the X chromosome using *Peddy*[4]. A sex check procedure compared genetically imputed sex with recorded sex on phenotype information. A total of 15 individuals were excluded due to their inferred sex not matching their recorded sex.

Relatedness

The PC-Relate method[5], implemented in *Hail*, was used to assess genetic relatedness between all samples. First, pairwise kinship coefficients were estimated between all pairs of samples using LD-pruned SNPs (max $r^2 < 0.5$) from a set of high-quality common variants (MAF > 5%). Next, the pairwise kinship coefficient was used to identify related individuals with a second-degree or closer relationship. Once such a relationship was inferred, the sample with a higher sequencing quality and more phenotypical information from each pair was retained for further analyses. This procedure eliminated 89 samples.

Hard Filters

Overall sample quality control was assessed using *Hail's* sample_qc function to generate sample metrics from the raw variant calls. Several metrics were assessed, and hard-call filters were derived to exclude low-quality samples and individual outliers. Samples were required to have call rates above 0.9, and individuals above or below 3 SD from the sample mean for each of the following were removed: number of SNPs (nSNPs), heterozygous-homozygous call ratio (rHetHomVar), number of singleton calls (nSingleton), transition-transversion ratio (rTiTv), and insertion-deletion (rInsertionDeletion). The hard filter QC removed 75 individuals who were outliers for one or more of the above metrics (**Supplementary Figure 1**).

Ancestry prediction and principal components estimation

*Peddy* was also used to infer the biogeographical ancestry of CLOZUK2 samples using Principal Components Analysis (PCA) and a support vector machine guided by samples of known ancestry from the thousand genomes project (1KG)[6]. The majority of individuals (99.5%) were classified by the algorithm as European, with only three samples classified as American and 9 as Admixed/Other. In order to avoid population stratification, a potential problem in rare variant studies, we excluded samples falling 4 SD out of the mean of PC1 and PC2 for European samples (**Supplementary Figure 2**). This procedure removed 10 individuals, of which 9 had been classified as European.

We then used the same subset of high-quality common variants (MAF > 5%) used to estimate relatedness to calculate 10 genetic principal components using *Hail's* Hardy-Weinberg-normalized PCA method (hl.hwe_normalized_pca). These PCs were later used as covariates in all regression analysis.

Summary of sample exclusions

Across all sample QC measures (exome coverage and quality, sex-check, ancestry, relatedness, and hard filters), we excluded 343 cases, and 2062 individuals were kept in the final sample and taken forward in our analysis.

Variant annotation

After extensive quality control, variants were annotated using the Ensembl Variant Effect Predictor v102[14] and CADD v1.6[7, 8] in *Hail*. Variants annotated as stop-gain, frameshift or splice donor/acceptor variants were grouped into the protein truncating variants class (PTVs). In addition, missense variants with CADD PHRED-score ≥ 20 were considered putatively damaging missense variants and included in the missense variant analyses. Variants included in each of those classes (i.e., PTVs or damaging missense) and presenting at a minor allele frequency (MAF) lower than 1% in both the filtered CLOZUK2 sample and the European subset of gnomAD v2.1.1[9] controls ('controls_nfe') were retained for further analyses.

#### Supplementary Results

##### Generalised linear mixed-effect model (GLMM) regression

The implementation of GLMMs in *glmmTMB*[10] allows for, in addition to standard mixed-effect models with fixed and random effect terms, fitting a wider class of "distributional regression" models[11]. These flexible models can be used to estimate the effects of predictors for the conditional mean of the outcome (sometimes referred to as "location") and/or its variance ("dispersion" or "scale"). This can increase the power of regression approaches by reducing the overall unaccounted residual ("error") variance, part of which can be captured by random effect terms[12] in GLMMs, as well as increasing the precision of estimates derived from fixed-effect terms[13]. Taking advantage of this feature of *glmmTMB*, we included predictors previously shown to influence the within-person variance of clozapine plasma concentrations[14, 15] in all our regression models: Clozapine dose, the time between the last clozapine dose and blood sample collection (TDS), sex, age, and age$^2$. **Table 2** in the main text shows the results for the PharmaADME core full regression model, with the estimated effect sizes and corresponding summary statistics for predictors of the mean and variance of clozapine levels. The inclusion of these predictors improved the fit of our models evidenced by a reduction in the RMSE (Root Mean Squared Error) statistic from 164.3 in the standard mixed-effect models to 163.8 in the model including both mean and variance parameters.

Further to our main analyses, we tested if the presence of rare damaging variants in the PharmaADME core genes could simultaneously impact the mean and variance of clozapine plasma concentrations, as carriers of these variants might have a larger within-subject variability due to their atypical metabolism. We indeed found evidence of this, but only for those carrying PTVs in the PharmaADME core list (**Supplementary Table 7**). However, these results should be evaluated cautiously as testing for variance predictors in a mixed-effect model setting is likely less powered than testing for predictors of the mean[13, 16], and only a small subset of individuals carried those variants in our sample (n = 73). With this consideration, PTV carriers being more likely to have larger variability in clozapine levels further suggests that the presence of these variants impacts clozapine metabolism and supports the utility of therapeutic drug monitoring (TDM) approaches for personalising clozapine prescriptions while rare genetic variation becomes a part of pre-emptive pharmacogenomic testing.

##### Ordinal mixed-effect model regression

We also aimed to assess whether the differences in metabolism observed in individuals carrying rare damaging variants in the PharmaADME core list would affect their probability of having clozapine plasma concentrations below (< 350 ng/mL), within (350–600 ng/mL), or above the therapeutic range (> 600 ng/mL). Regression models for the bins of clozapine plasma concentrations were fitted with ordinal GLMMs assuming a cumulative link function for the bins of clozapine plasma concentrations via the *ordinal* R package[17]. This regression included the same fixed-effect and random-effect covariates from the models described above. We found that individuals carrying at least one rare damaging variant in PharmaADME core genes were 30% less likely than non-carriers to reach clozapine concentration within or above the therapeutic range (OR= 0.694; SE= 0.145; P= 0.012). **Supplementary Figure 5** shows that those carrying rare damaging alleles were at increased likelihood of presenting subtherapeutic clozapine concentrations, requiring doses of 275 mg/day to reach the therapeutic interval with at least 50% probability (50.43%; SE 2.48%), compared to individuals without those variants achieving the same outcome at 225 mg/day (50.44%; SE 2.06%).

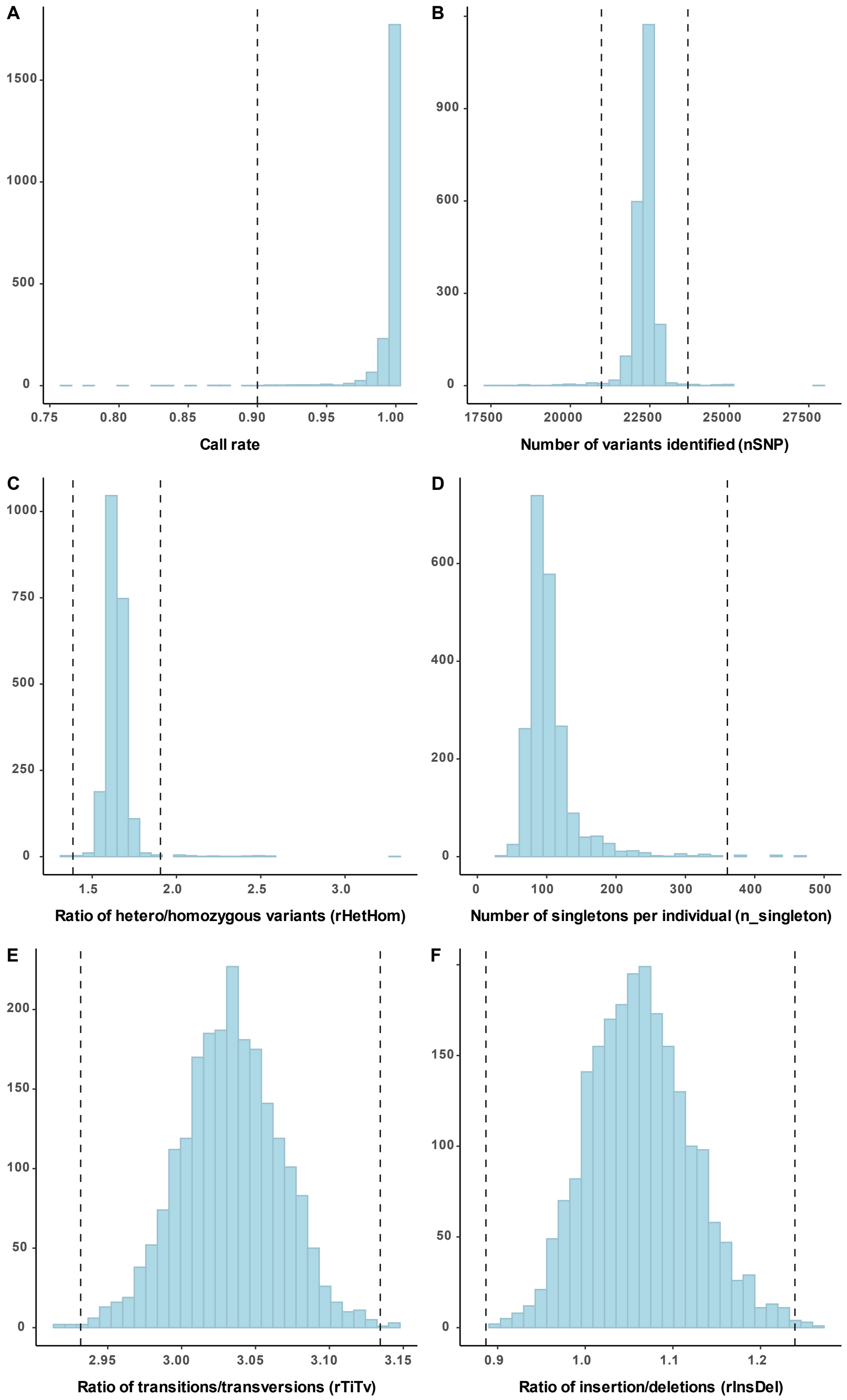

Supplementary Figure 1: Histograms showing the distributions of sample metrics used in the hard filter QC. The dotted lines indicate the thresholds used to filter samples from the analysis.

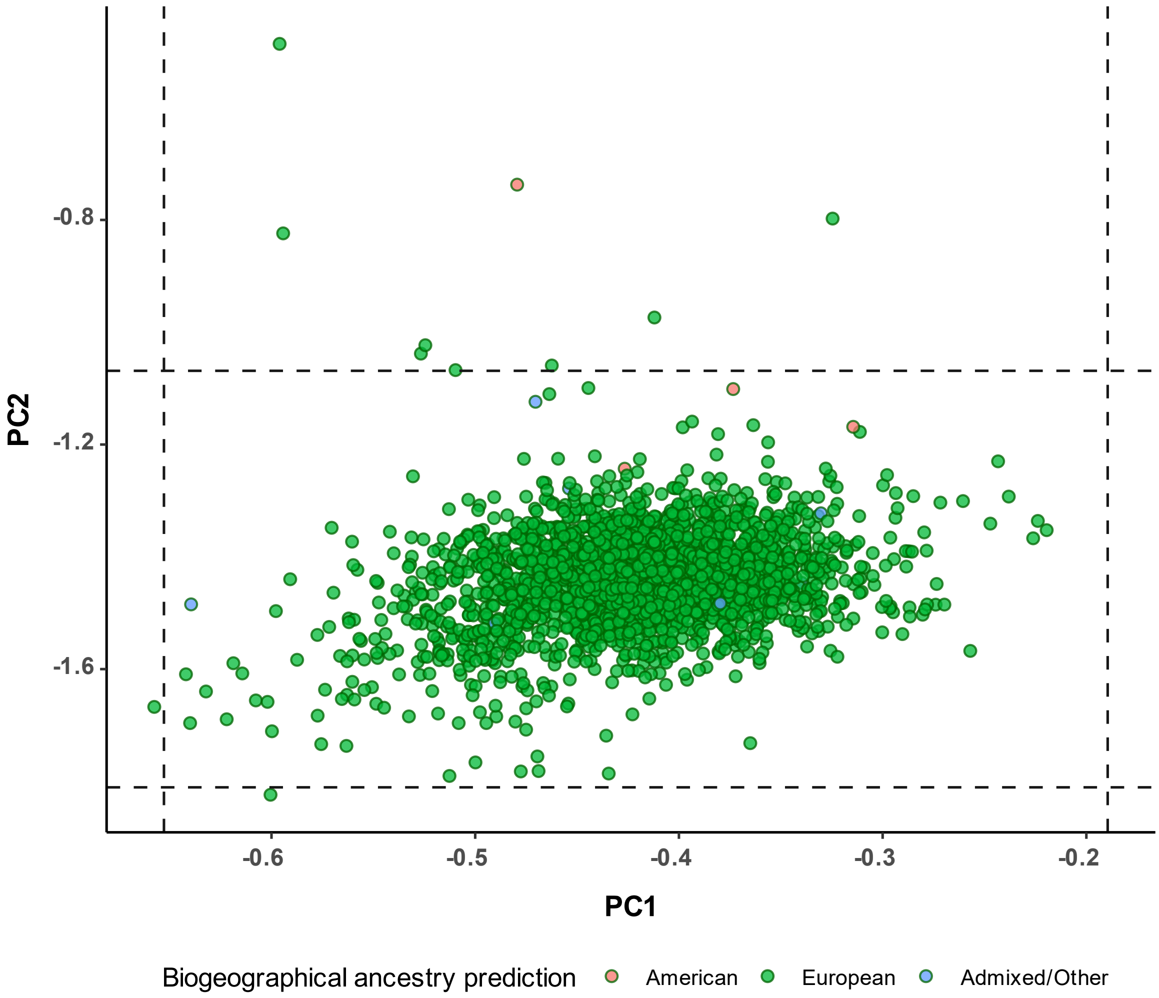

Supplementary Figure 2: Principal components analysis of CLOZUK2 samples. The dotted lines show the thresholds used to exclude samples to avoid population stratification. Colours represent the individual's biogeographical ancestry prediction derived in *Peddy*.

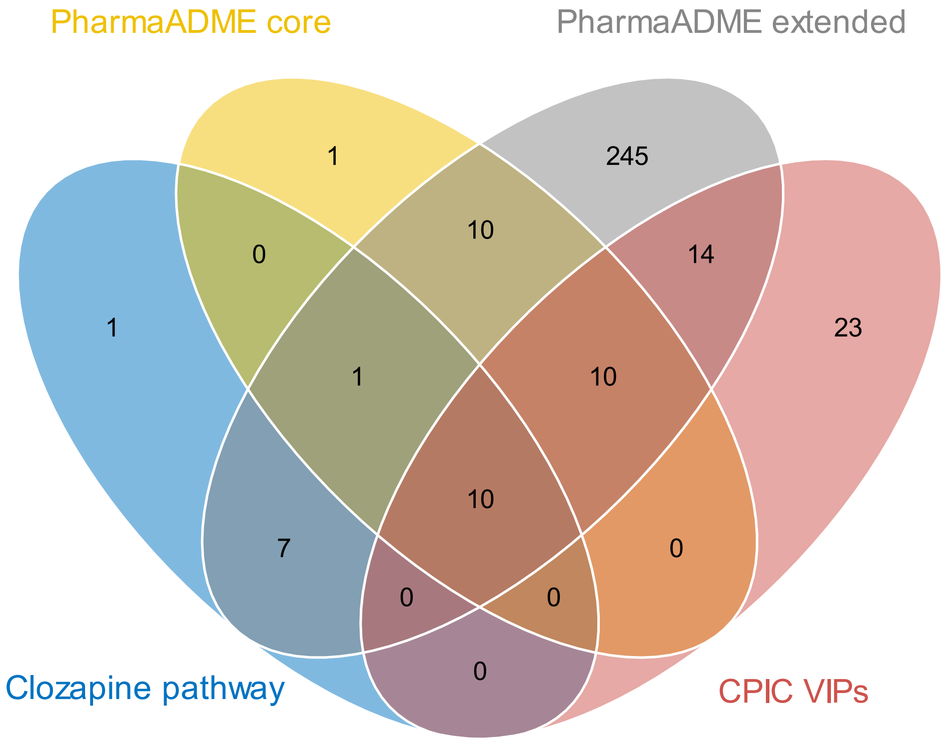

Supplementary Figure 3: Venn diagram showing the intersection of all gene sets analysed.

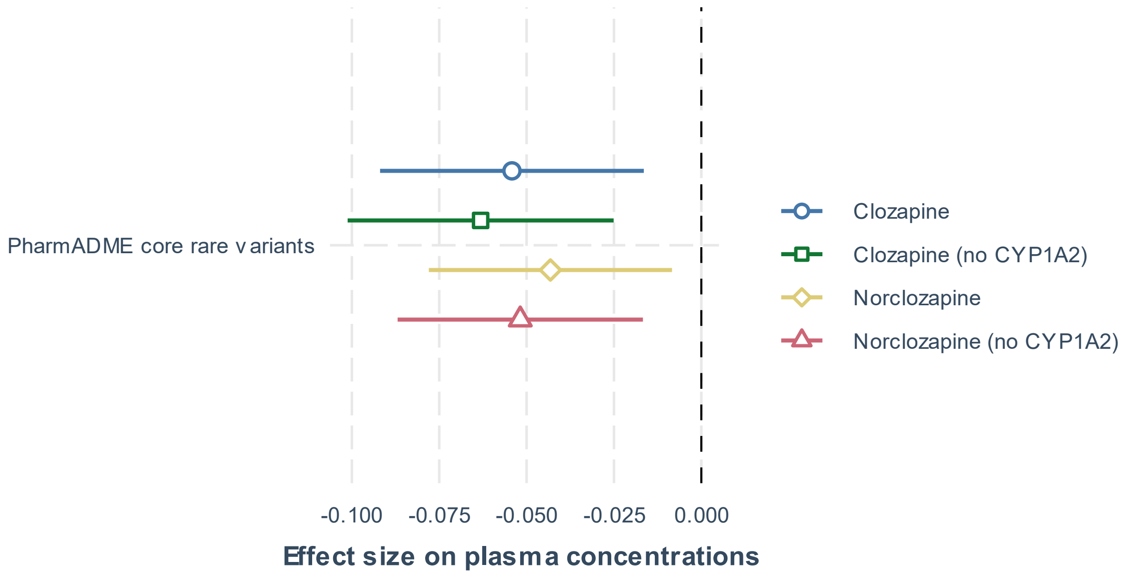

Supplementary Figure 4: Differences in the effect size of rare damaging variants in PharmaADME core set list on clozapine metabolism phenotypes when including or removing *CYP1A2* variants.

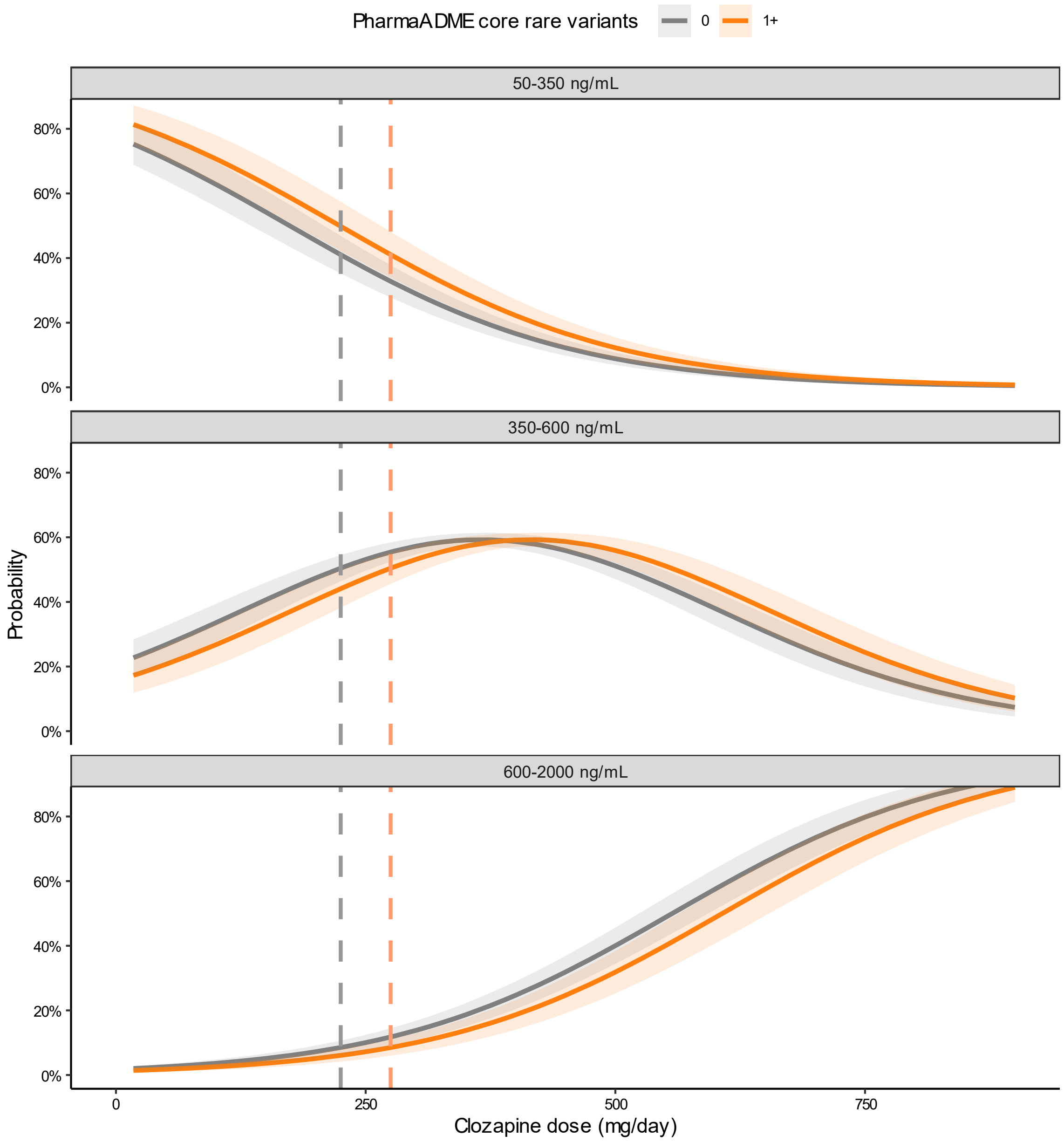

**Supplementary Figure 5**: Marginal effects derived from an ordinal mixed model regression for the probability of achieving clozapine concentrations in or out of the therapeutic range for individuals carrying none or at least one rare damaging variant in the PharmaADME core gene list. Shaded areas indicate a 95% confidence interval. Vertical dashed lines highlight the doses individuals require to reach the therapeutic range with a 50% probability, given their rare variant burden.
